# Reasons underlying the intention to vaccinate children aged 5-11 against COVID-19: A cross-sectional study of parents in Israel, November 2021

**DOI:** 10.1101/2022.03.03.22271793

**Authors:** Nicole G. Morozov, Amiel A. Dror, Amani Daoud, Netanel Eisenbach, Edward Kaykov, Masad Barhoum, Tsvi Sheleg, Eyal Sela, Michael Edelstein

## Abstract

Vaccination is a key tool to mitigate impacts of the COVID-19 pandemic. In Israel, COVID-19 vaccines became available to adults in December 2020 and to 5–11-year-old children in November 2021. Ahead of the vaccine roll-out in children, we aimed to determine whether surveyed parents intended to vaccinate their children and describe reasons for their intentions. We collected information on parental socio-demographic characteristics, COVID-19 vaccine history, intention to vaccinate their children against COVID-19, and reasons for parental decisions using an anonymous online survey. We identified associations between parental characteristics and plans to vaccinate children using a logistic regression model and described reasons for intentions to vaccinate or not. Parental non-vaccination and having experienced major vaccination side effects were strongly associated with non-intention to vaccinate their children (OR 0.09 and 0.18 respectively, p<0.001). Parents who were younger, lived in the socio-economically deprived periphery, and belonged to the Arab population had lower intentions to vaccinate their children. Reasons for non-intention to vaccinate included concerns about vaccine safety and efficacy (53%, 95%CI 50-56) and the belief that COVID-19 is a mild disease (73%, 95%CI 73-79), while a frequent motive for vaccination was the return to normal social and educational life (89%, 95%CI 87-91). Understanding rationales for COVID-19 vaccine rejection or acceptance, as well as parental demographic data, can pave the way for intentional educational campaigns to encourage not only vaccination against COVID-19, but also regular childhood vaccine programming.

**Highlights:**

- Parental intention to vaccinate children aged 5-11 is much lower than vaccine coverage in parental age groups
- Being unvaccinated and having experienced side effects following vaccination were the greatest negative predictors in parents of intention to vaccinate their children
- Parents were more likely to accept a COVID-19 vaccine for their children to allow them to return to daily social life and to ensure economic security in the family
- Parents were more likely to reject a COVID-19 vaccination for health reasons such as safety concerns or due the belief that COVID-19 was a mild disease in children

## Introduction

The rapid, global spread of COVID-19, an infection caused by the SARS-CoV-2 virus, spurred the expeditious development of several vaccines and the eventual implementation of vaccine programs at record speeds. In Israel, COVID-19 vaccines became available in December 2020 for adults and eligibility gradually extended to younger ages. Teenagers aged 12-15 became eligible in June 2021 and children aged 5-11 in November 2021. Most high-income countries now offer teenage vaccination, but vaccination of younger children is less common despite evidence of its safety and effectiveness ^1^. Outside of Israel, the United States’ (USA) Centers for Disease Control and Prevention (CDC) recommends vaccination in the 5-11 age group while the European Centre for Disease prevention and Control has also recommended considering universal vaccination in this demographic^2, 3^.

The short period between the pandemic’s onset and vaccine availability has added additional challenges for vaccine acceptance among the public ^4^. Almost two years after the onset of the pandemic and one year after the first administration of COVID-19 vaccines in adults, the USA’s Food and Drug Administration (FDA) approved the Pfizer/BioNTech mRNA vaccine for 5-to 11-year-old children ^1, 5^. Studies conducted in the UK, USA, and Canada suggest that many parents remain concerned about the vaccine’s side effects and safety, leading parents to be hesitant to administer the COVID-19 vaccine to their children ^6-8^. While new research has examined the reasons why parents would be motivated or hesitant to vaccinate their children in Israel, ours is one of the first to examine attitudes of parents to children aged 5-11.

Demographic criteria which have been associated with higher rates of vaccine refusal for children include younger parental age, non-white or minority ethnicity, lower education level, history of vaccination non-compliance to recommended childhood vaccinations, and lower income in countries such as England. Canada, and Brazil ^7, 9-11^. Parents concerned about future finances were more likely to accept a vaccination for their offspring ^11^. Larson et al. utilized a four-question survey across emergency departments in 2016 to uncover the differing beliefs towards vaccine safety across 67 countries, finding that vaccine safety skepticism relied on both the specific vaccine type and the country itself ^12^. Compared with most high-income countries, Israel typically achieves high coverage for routine childhood vaccines. The 2020 national coverage for the first MMR dose among 24-month-olds was 99% in Israel vs 91% in the United States and in the UK ^13^. Of note, Arab Israelis typically achieve higher coverage for routine childhood vaccines compared with the general population ^14^. It is, therefore, important to understand and document attitudes to COVID-19 vaccinations due to their implications for planning public health interventions.

We aimed to investigate the intentions of surveyed parents in Israel to vaccinate their 5-11-year-old children against COVID-19 and the underlying reasons for their intentions. We sought to determine what common reasons parents listed for accepting or rejecting vaccinating their children. Our survey was initiated two weeks before children in this age group became eligible for COVID-19 vaccination.

## Methods

### Study design

This study was designed as a cross sectional survey to understand the willingness of surveyed parents of children 5-11 years old to vaccinate their children against COVID-19 and the reasons for their acceptance or refusal of the vaccine using an online questionnaire in Hebrew and Arabic. The survey was distributed through social media channels (Facebook and Twitter). Participants voluntarily participated in the survey. Eligibility criteria for inclusion in this study included whether the participant is a parent and if the parent has a child between the ages of five and 11. Appendix 1 provides an example in English of the study with the goal of the survey to determine eligibility for inclusion into our study, to obtain demographic data of the participant, and to establish the participant’s intention to vaccinate their 5–11-year-old children. Informed consent was requested on the introductory page of the online questionnaire, prior to survey enrollment. The anonymous web-based survey followed the American Association for Public Opinion Research (AAPOR) reporting guidelines ^15^. The survey was anonymous and no identifiable information such as name, date of birth, or identification was collected. Participants were permitted to terminate participation at will. Approval of this study was granted by the Research Ethics Committee of the Galilee Medical Center. The questionnaire was designed and distributed using Qualtrics (Qualtrics, Provo, UT, United States of America).

The survey was distributed between November 1-14, 2021, spanning over 14 days and terminating one day before the planned opening of COVID-19 vaccinations to 5-11-year-old children in Israel. The outreach to popular social media channels in Israel was conducted through Facebook, Instagram, and Twitter advertisement platforms. To encourage participation from a wide range of individuals representing Israeli society, we used advertisement sets in both Arabic and Hebrew and tailored to males and females. We used the social media channel’s advertising algorithm to target parents of young children. Only respondents who were older than 18 years, were parents of children aged 5-11, lived in Israel, and who completed the survey were included in this study. The survey was distributed evenly in all geographic regions of Israel. The advertisement invitation to participate in our survey included a direct callout with the phrase “Can you help us to understand parent intention to vaccinate children for COVID-19” followed by “Voluntary survey to study parents’ intention to vaccinate children aged 5-11 for COVID-19”. The impression segmentation and click conversion rate are described in Figure 1.

**Figure 1:**
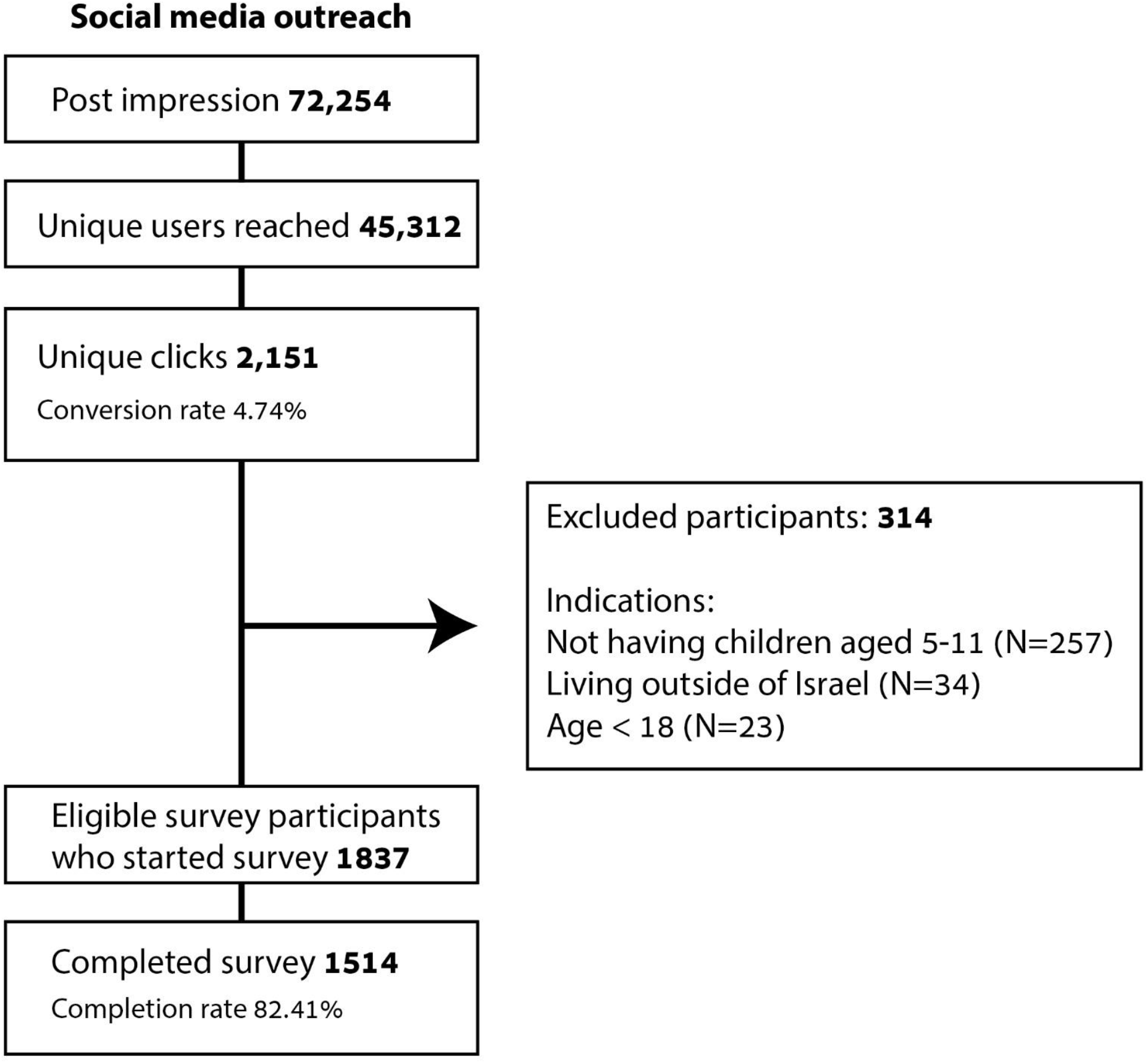
Social media outreach results, including initial post impressions, attrition, exclusion criteria, and conversion rate to clicks.

### Survey Instrument

The survey had two sections: The first section included nine questions about socio-demographic characteristics of the participating parents including parental age, gender, area of residence, household composition, number of children, ethnicity, parental education, parental COVID-19 vaccination status, and self-reported side effects from the vaccine (major, mild, or no symptoms), and one question on intention to vaccinate their child(ren) against COVID-19.

In the second section, responders who indicated they were not intending to vaccinate their children against COVID-19 were asked to state their reasons from a list of predefined explanations, and those who indicated intention to vaccinate were also asked to affirm why from a list of potential reasons. A free text answer was also provided.

The full questionnaire is available in Appendix 1. The estimated answering time for completing the survey was under two minutes.

### Statistical analysis

We calculated the proportion of parents intending to vaccinate their children together with 95% confidence intervals, overall and in each stratum. We calculated that in order to detect a 5% difference in response between vaccination intenders and non-intenders with an alpha of 0.05 and a power of 80%, 1531 participants were required. We compared parents who intended to vaccinate their children to those who did not in terms of all socio-demographic factors recorded in the questionnaire using a univariate logistic regression model. To determine the extent to which each factor was an independent predictor, we conducted a multivariate logistic regression analysis adjusting for all other factors found to be statistically significant (at the alpha=0.05 level) in the univariate analysis. We also calculated the proportion of parents giving each reason for intending or not intending to vaccinate together with 95% confidence intervals.

Data were prepared using Microsoft Excel and analyzed using STATA 15.

## Results

We received 1837 responses to our survey, published online between November 1-14, 2021. Of those, 1514 (82.4%) completed the survey in its entirety. The distribution of responders according to age, sex, ethnicity, education level, and other demographic criteria is described in Table 1. In our study parental self-reported ethnicity was 70.7% Jewish, 25.5% Arab, and 3.8% “other”. In the general population these proportions, as of December 2021 were, 73.9%, 21.1% is Arab, and 5.0% respectively.^16^ Overall intention to vaccinate was 44% (95% CI 41.3-45.9).

Female parents and parents above the age of 35 were more likely to vaccinate than male parents and parents aged 35 and younger (47% vs. 40%, p<0.002; 45% vs. 39%, p<0.02, Table 1). Compared with non-Arab Israeli parents, Israeli Arab parents reported a lower intention to vaccinate their children (34% vs. 47%, p<0.001, Table 1). Other parental factors significantly associated with a lower intention to vaccinate included having a single child, living in the periphery (Northern or Southern, socioeconomically depressed regions of the country), having received fewer than three doses of a COVID-19 vaccine and having experienced major side effects of COVID-19 vaccination. After adjusting for other factors, being male, living in the periphery, belonging to the Arab population, reception of fewer than three doses of the COVID-19 vaccine and having experienced major side effects following COVID-19 vaccination all remained significantly associated with a lower intention to vaccinate children (Table 1). Being completely unvaccinated and having suffered major side effects were associated with the largest reduction in intention to vaccinate children (−91% and -81% respectively, p<0.001).

Of 801 parents intending to vaccinate their children, 93% were COVID-19 vaccinated at least once. Intention to vaccinate their children increased with the number of doses the parents had received; while only 13% of unvaccinated parents intended to vaccinate their children against COVID-19, 32% of parents vaccinated with one dose, 48% of parents vaccinated with two doses, and 62% of parents vaccinated with three doses were willing to vaccinate their children (p<0.001, Table 1).

The most often cited reason for choosing to vaccinate their children was to allow their children to return to everyday social, educational, and financial life, followed by protecting other family members from COVID-19 infection (89 and 77.9% respectively, Table 2). The protection of the children against viral COVID-19 infection followed by vaccine safety were the least often mentioned explanations for parents who intended to vaccinate their children (37.9%).

The most frequently mentioned explanation for not vaccinating their children is the belief that COVID-19 leads to mild or asymptomatic disease in children (76.1%, Table 2). Concerns of the parents regarding vaccine safety were ordered second in their consideration not to vaccinate their children (53.1%, Table 2).

## Discussion

Routine childhood vaccination offers the prospect of protection against infectious diseases. In most settings, there is no ongoing endemic transmission of poliovirus, measles, or diphtheria infections due to effective global vaccination campaigns ^17-19^. However, COVID-19 is unique in several ways: first, the pandemic is continuing to develop and therefore vaccine protection is against an immediate and tangible threat; second, the volume and availability of information around COVID-19 vaccines, offering arguments both for and against vaccination with very few verification pathways for parents to assess the accuracy of the information they are accessing, is an unprecedent phenomenon; third, the speed at which COVID-19 vaccines were developed have raised some concerns about safety among the general public; fourth, the shifting understanding that the purpose of the COVID-19 vaccines, including mRNA vaccines, is not to prevent infection but rather to decrease the likelihood of severe adverse outcomes ^20, 21^.

Existing research has established several factors which may influence vaccine acceptance, including the type of vaccine, home country of the individual, religiosity, age, and other socio-demographic criteria ^7, 9-11^. These links were studied also in association with the global outbreak of the H1N1 influenza pandemic of 2009-2010. According to a World Health Organization systematic review report in 2016 of attitudes towards vaccination during this particular influenza pandemic, parents of children aged 6-59 months were less likely to accept a vaccine for their children if the parents were younger and had a lower educational achievement ^22^. The most cited reasons for hesitancy, during the 2009-10 influenza pandemic, included a decreased perception of benefit from the influenza vaccine, decreased perception of danger from the virus, and difficulties with ease of access to vaccination ^22^. Similarly, we found that the most commonly held reason by surveyed parents to not vaccinate their children is the belief that the disease is mild or asymptomatic in children.

The COVID-19 pandemic has seen the digital spread of information, misinformation, and disinformation on social media platforms on an unprecedented scale ^23^. Mistrust in government and international health organizations has been suggested as a potential contributor to hesitancy to follow government and international health guidelines, as fewer than 30% of individuals opted to search for their own government social media publications for vaccine information and guidance ^24^. Demographic data from social media sites shows that the largest user age groups of social media sites such as Facebook, Instagram, and YouTube are ages up to 35 years old ^25^. Our results indicate that parents younger than 35, the greatest social media users, were less likely to vaccinate their children.

Of additional importance in the discussion of COVID-19 vaccine hesitancy is whether an individual has experienced adverse effects attributed to the vaccine. Parental experience was clearly a key factor in the decision, with unvaccinated, surveyed parents and those who experienced perceived severe side effects very unlikely to vaccinate their children. Vaccine hesitant individuals who experienced an adverse effect attributed to a vaccine did receive the vaccine, indicating a likely initial willingness to be vaccinated but subsequent hesitancy based on experience. Acknowledging these experiences while correcting potential factual misconceptions regarding vaccine safety may be a technique to assuage concerned parents, as suggested by a clinical report from The American Academy of Pediatrics ^26^. Healthcare providers may be better prepared to repair their trust in vaccines in the future, thus paving the way for parents to have greater trust in vaccines and subsequent willingness to vaccinate their children.

We found that the intention to vaccinate children aged 5-11 was low, overall. Less than half of surveyed parents intended to vaccinate their 5-11 children, and as of February 2022 just over 20% had vaccinated their offspring ^27^. Even among parents who had received three doses of the vaccine, intention to vaccinate was only 62%. This is consistent with experience from other countries where vaccinated parents did not intend to vaccination their children ^28^. Data from a separate study, available online but not yet in a peer-reviewed publication, conducted at the same time showed that, similar to our results, only 37% of parents intended to vaccinate their children^29^. An Israeli study conducted prior to the initiation of ours found that Israeli parents of children of any age, with a bachelor’s degree, were found to have higher rates of vaccination ^30^. In contrast, we found that higher educational attainment did not reflect higher vaccination levels; the differences in these findings may be related to the age of parents. While the parents in our study had at least one child in the 5-11-year-old range, the parents in Heller et al.’s investigation were not necessarily parents to children in that age bracket.

Our study has highlighted that the main reasons for intending to vaccinate children were not biomedical. Key factors within Israeli society associated with an increased intention to vaccinate children against COVID-19 include the parent’s desire for the child to return to normal social and educational life along with secondary motives such as the parent’s desire to maintain job security. Our findings suggest that, for COVID-19, the traditional perception of vaccination benefits such as protection against severe illness has been superseded by indirect benefits such as returning to regular societal life and education institutions, as well as assuring financial resilience for the family. While this finding is not surprising considering the severe disruption to normal life caused by the COVID-19 pandemic, it is not clear whether this perception of the societal utility of vaccines will continue to impact the perception of other vaccines beyond the pandemic.

The main drivers against intention to vaccinate are more biomedical, encompassing beliefs that COVID-19 causes a mild disease course in children and concerns about vaccine safety. Heller et al. found vaccine hesitancy to be greater among respondents who did not believe COVID-19 vaccines are safe, with those who believe it is safe having higher rates of vaccination ^30^. Interestingly, the results of their study, which was able to demonstrate longitudinal attitudes towards COVID-19 and vaccination, indicated a decreasing fear of COVID-19 and a greater distrust in vaccines ^30^. Despite that finding, Heller et al. discovered that a greater proportion of respondents who had initially been worried about vaccine safety were vaccinated over the study’s time period, a finding which may suggest a change in policy and incentives or education about vaccines ^30^. These findings have implications for health promotion and communication campaigns; ensuring parents’ confidence in the safety of the vaccine and highlighting the sociocultural benefits of vaccination may be key in guiding communities to higher levels of vaccine uptake.

Groups who may be targets for intersectional outreach include minorities, parents with a lower educational level, and families in lower financial brackets as these parents have been highlighted as more likely to be vaccine hesitant ^10, 31^. Our research found that Arab parents were less likely to vaccinate their children as compared to other parents, consistent with research finding a higher rate of COVID-19 vaccine hesitancy among minority communities in other countries, as well as in Israel’s Arab and Ultra-Orthodox communities ^7, 32, 33^.

Strategies to reduce vaccine hesitancy have been discussed in other literature. Financial or other incentivizing techniques for parents have been shown to increase vaccine uptake among their children in certain communities such as in Nicaragua and India ^34^. A major motivator for vaccine uptake in our study is a desire to return to normal social and educational life in Israel, which is an incentive in and of itself. Use of technology and social marketing has also been shown to increase vaccine acceptance for HPV and MMR vaccines ^34^. Digital health education interventions specific to parental concerns, such as about vaccine technologies, may be key to targeting certain parental groups.

The key limitation of our study is inherent to its design and concerns the self -selected nature of the sample.. Parents who do not use social media channels, and therefore would not have been exposed to this study, may not have been appropriately represented. Although use of social media use is widespread in parental age groups in Israel, certain groups, for example the Ultra-Orthodox, may be underrepresent due to lower smartphone and Internet penetration ^35^. Because the survey is entirely anonymous, comparisons between the sample and the general population are difficult. Nevertheless, the ethnicity distribution of respondents was comparable to the general population. Another potential selection bias is that individuals who have stronger opinions either for or against vaccinating their children for COVID-19 may be more likely to complete a questionnaire about the topic. It is also theoretically possible for individuals to have completed the study more than once given that identifying participant information was not collected. It must be noted that the age, sex, ethnicity, and geographical distribution of participants suggest that all segments of the population were represented.

It is not clear to what extent our findings are generalizable to other countries. Globally, there are wide discrepancies in vaccine uptake from one country to another with some countries enjoying remarkably low levels of vaccine hesitancy among parents ^9, 11^. Understanding what factors enable childhood COVID-19 vaccination in countries with high parental intent and transferring this knowledge to other contexts could help countries struggling with COVID-19 childhood vaccination programs. Because a growing number of individuals, including parents, are seeking health and vaccine information online or through peers, the availability of trustworthy sources and early educational campaigns cannot be overstated ^36^.

## Conclusion

The availability of COVID-19 vaccines and licensing of these vaccinations for use in children aged five to 11 is not a guarantee that parents will inoculate their children. Our study showed that intention to vaccinate 5- to 11-year-old children was low even among fully vaccinated parents, with socio-cultural factors, as well as vaccination experience in parents, being associated with intention to vaccinate their children. Our study showed that indirect benefits of vaccination such as returning to regular societal activities, including educational institutions and personal economic stability, were bigger motivators than direct protection against the virus, while the most common reasons for intending not to vaccinate children against COVID-19 were classical biomedical concerns such disease severity and vaccine safety. These findings are important for health authorities to shape educational and communication campaigns: focusing on the message that vaccines protect against COVID-19 may not be as effective as messages stating that vaccines will allow children to go to school and adults to work. Different messaging campaigns should be implemented and evaluated to test this possibility.

## Data Availability

All data produced in the present study are available upon reasonable request to the authors

## Attestation

All authors attest they meet the ICMJE criteria for authorship

## Data availability statement

Data is available upon request from the corresponding author.

