## Supplementary material for "Reasons underlying the intention to vaccinate children aged 5-11 against COVID-19: A cross-sectional study of parents in Israel, November 2021": Table 1

Table 1. Parental socio-demographic background and vaccination history and intention to vaccinate

| | | Number of respondents | Number intending to vaccinate their 5–11-year-old child | Percent intending to vaccinate their 5-11-year-old child | 95% CI | Unadjusted OR | Unadjusted OR 95%CI | P value | Adjusted OR* | Adjusted OR 95%CI | P value |
|---|---|---|---|---|---|---|---|---|---|---|---|
| **Total** | | 1837 | 801 | 43.6 | 41.3-45.9 | - | - | | - | - | |
| **Parent sex** | Female | 891 | 422 | 47.4 | 44-50.7 | baseline | | | | | |
| | Male | 946 | 379 | 40.1 | 36.9-43.2 | 0.74 | 0.62-0.89 | 0.002 | 0.75 | 0.6-0.94 | 0.01 |
| **Parent age** | >35 | 1309 | 594 | 45.4 | 42.6-48.1 | baseline | | | | | |
| | ≤35 | 528 | 207 | 39.2 | 35-43.5 | 0.78 | 0.74-0.93 | 0.02 | 0.85 | 0.67-1.1 | 0.2 |
| **Number of children** | One child | 421 | 161 | 38.2 | 33.6-43.1 | 0.75 | 0.6-0.94 | 0.01 | 0.74 | 0.57-0.96 | 0.02 |
| | > one child | 1416 | 640 | 45.2 | 42.6-47.8 | baseline | | | | | |
| **Household composition** | Two parents | 1568 | 669 | 42.7 | 40.2-45.1 | 0.77 | 0.6-1 | 0.05 | 0.8 | 0.59-1.1 | 0.2 |
| | Not two parents | 269 | 132 | 49.1 | 42.9-55.2 | baseline | | | | | |
| **Parent education level** | High school or under | 379 | 161 | 42.5 | 37.4-47.6 | baseline | | | | | |
| | Bachelor's degree | 1155 | 525 | 45.5 | 42.5-48.4 | 1.13 | 0.89-1.4 | 0.3 | - | - | - |
| | Higher degree | 303 | 115 | 38.0 | 32.5-43.7 | 0.83 | 0.61-1.13 | 0.2 | - | - | - |
| **Resident area** | Center | 1103 | 508 | 46.1 | 43.1-49 | baseline | | | | | |
| | Periphery | 734 | 293 | 39.9 | 36.3-43.6 | 0.78 | 0.64-0.94 | 0.01 | 0.73 | 0.59-0.92 | 0.006 |
| **Parent ethnicity** | Non-Arab Israeli | 1298 | 611 | 47.1 | 44.3-49.8 | baseline | | | | | |
| | Arab Israeli | 469 | 159 | 33.9 | 29.6-38.4 | 0.58 | 0.46-0.72 | <0.001 | 0.65 | 0.51-0.84 | 0.001 |
| | Other | 70 | 31 | 44.3 | 32.4-56.7 | 0.89 | 0.55-1.45 | 0.6 | 1.3 | 0.71-2.3 | 0.4 |
| **Parent COVID-19 vaccination status** | 3 vaccine doses | 578 | 357 | 61.8 | 57.7-65.7 | baseline | | | | | |
| | 2 vaccine doses | 728 | 351 | 48.2 | 44.5-51.9 | 0.57 | 0.46-0.72 | <0.001 | 0.61 | 0.49-0.77 | <0.001 |
| | 1 vaccine doses | 125 | 40 | 32.0 | 23.9-40.9 | 0.29 | 0.19-0.43 | <0.001 | 0.29 | 0.19-0.45 | <0.001 |
| | Not vaccinated | 406 | 53 | 13.1 | 9.9-16.7 | 0.09 | 0.07-0.13 | <0.001 | 0.09 | 0.07-0.13 | <0.001 |
| **Parent COVID-19 vaccination side effects** | Yes: major | 112 | 22 | 19.6 | 12.7-28.2 | 0.18 | 0.11-0.29 | <0.001 | 0.19 | 0.11-0.31 | <0.001 |
| | Yes: minor | 546 | 279 | 51.1 | 46.8-55.3 | 0.76 | 0.61-0.95 | 0.02 | 0.76 | 0.60-0.95 | 0.02 |
| | No | 773 | 447 | 57.8 | 54.2-61.3 | baseline | | | | | |
| | Missing data | 406 | | | | | | | | | |
| | For variable significance in the univariate analysis, adjusted for other significant variables | | | | | | | | | | |
