## Supplementary material for "Reasons underlying the intention to vaccinate children aged 5-11 against COVID-19: A cross-sectional study of parents in Israel, November 2021": Table 2

Table 2. Stated reasons given by parents for intention or refusal to vaccinate their children, Israel, November 2021

| | | n | % | 95% ci |
|---|---|---|---|---|
| I want to vaccinate my child…(n=801) | to return to normal life and activities | 713 | 89 | 86.6- 91.1 |
| | to loss and secure income | 624 | 77.9 | 74.8-80.7 |
| | to protect others | 560 | 69.9 | 66.6-73.1 |
| | to protect my child | 448 | 55.9 | 52.4-59.4 |
| | because the vaccine is safe | 304 | 37.9 | 34.6-41.4 |
| I don't want to vaccinate my child… (n=1036) | because COVID-19 is a mild illness | 788 | 76.1 | 73.3-78.6 |
| | because of vaccine safety concerns | 550 | 53.1 | 50-56.2 |
| | because I don't trust the government | 435 | 42 | 39-45 |
| | because I don't trust the Ministry of Health | 217 | 21 | 18.5-23.5 |
| | Because I think COVID-19 is a conspiracy | 145 | 14 | 11.9-16.2 |
