## Appendix 1 for "Reasons underlying the intention to vaccinate children aged 5-11 against COVID-19: A cross-sectional study of parents in Israel, November 2021"

### Survey of parental intention to vaccinate their child against COVID-19

(Among parents with children aged 5-11 years old – Israel, Nov 2021)

We have posted an online cross-sectional questionnaire and invited parents of children aged 5-11 years old to participate through social media channel (Facebook and Twitter). The invitation post was simultaneously shared on these platforms on November 14, 2021 and data was collected over the course of two weeks. The survey included 20 question written in Hebrew with a validated translation to Arabic performed following the translation protocol: (1) The original Hebrew survey (suppl table 1) was first distributed among 20 Hebrew speaking volunteers to assess the literacy of the questionnaire and to assure that the text or phrases are not offensive to participants; (2) the translation to Hebrew by native Arabic speaking Israeli Arabs; (3) re-translation from Arabic to Hebrew by three independent translators who speak Arabic and Hebrew at the level of fluency; (4) the three independent back-translations (Arabic to Hebrew) were examined for consistency, with non-matching translations followed the three translators agreeing on a common, suitable translation. Finally, the survey was distributed among 20 Hebrew speaking and 20 Arabic speaking volunteers to check the comprehensiveness and literacy of the questionnaire prior to formal distribution.

**Demographic Block**

---

1. Gender
☐ Male
☐ Female

2. Age
☐ ≤35
☐ >35

3. Children aged 5-11
☐ One child
☐ Two or more

4. Parents
☐ Two parents
☐ Other

5. Education
- ☐ Undergraduate
- ☐ Bachelor
- ☐ Higher degree

---

6. Resident
- ☐ Centre
- ☐ Tel Aviv
- ☐ Jerusalem
- ☐ Haifa
- ☐ North
- ☐ South

For analysis purposes, the residency areas were grouped accordingly:
Center = Centre, Tel Aviv, Jerusalem, and Haifa
Periphery = North and South

---

7. Ethnicity
- ☐ Arab Israeli
- ☐ Non-Arab Israeli
- ☐ Others

---

8. COVID-19 vaccine doses
- ☐ One vaccine
- ☐ Two vaccines
- ☐ Three vaccines
- ☐ None

---

9. Vaccine side effects
- ☐ Major
- ☐ Minor
- ☐ No side effect

---

10. Intention to vaccinate child
- ☐ Yes
- ☐ No

**End of Block**

---

**Start of Block:** *This section is presented to you since you responded that you are* ***willing to vaccinate your child***

Which, if any, of the following reasons for intention to vaccinate children against COVID-19 applies to you?

| Reasons to vaccinate my child | | |
|---|---|---|
| 11. Return to normal life and activities | ☐ Major reason | ☐ Not a reason |
| 12. Secure income and avoid job loss | ☐ Major reason | ☐ Not a reason |
| 13. Protect others | ☐ Major reason | ☐ Not a reason |
| 14. Protect child | ☐ Major reason | ☐ Not a reason |
| 15. Vaccine is safe | ☐ Major reason | ☐ Not a reason |
| Other: ____________________________________________ | | |

**End of Block**

---

**Start of Block:** *This section is presented to you since you responded that you are* ***not willing to vaccinate your child***

Which, if any, of the following reasons for intention to vaccinate children against COVID-19 applies to you?

| Reasons not to vaccinate my child | | |
|---|---|---|
| 16. Believe that COVID-19 leads to mild illness in children | ☐ Major reason | ☐ Not a reason |
| 17. Concerned about COVID-19 vaccine safety/side-effect for children | ☐ Major reason | ☐ Not a reason |
| 18. Distrust in government decisions for vaccinating children | ☐ Major reason | ☐ Not a reason |
| 19. Distrust in health ministry | ☐ Major reason | ☐ Not a reason |

20. Other: ______________________________________________
